# Diagnostic Utility of Electrophysiological Markers for Early and Differential Diagnosis of Alzheimer’s, Frontotemporal, and Lewy Body Dementias: A Systematic Review

**DOI:** 10.1101/2024.09.20.24314042

**Authors:** Stéphanie De Keulenaer, Sara Van Mossevelde, Tobi Van den Bossche, David Crosiers, Patrick Cras, Tommas Ellender, Rose Bruffaerts

## Abstract

**Background:** An early and accurate diagnosis is crucial to provide optimal patient care in neurodegenerative diseases. Although an EEG shows advantages in availability and cost compared to the current diagnostic tools, it is not routinely used in clinical practice. Previous reviews have either focused on single disease populations and/or solely on resting state EEG. To evaluate the utility of EEG for early diagnosis and differential diagnosis, we conducted a systematic review across Alzheimer’s disease (AD), Frontotemporal Dementia (FTD) and Lewy Body Dementia (DLB).

**Methods:** We searched databases Pubmed, Cochrane, Web of Science, and Scopus for articles published from 2000 to 2023 investigating resting-state and task-based EEG-markers in biomarker-proven AD, FTD and DLB.

**Results:** Our search yielded a total of 12010 studies, of which 71 papers were eligible: 34 on AD, 18 on DLB and 9 on FTD. Slowing of the frequency spectrum was a common observation across diseases, achieving excellent sensitivity in AD and DLB. Research on FTD was limited and with varying results in the discrimination from healthy controls, although connectivity analysis and microstates are promising avenues. In differential diagnosis, both spectral and connectivity metrics show encouraging results. Task-based EEG emerges as a promising tool in early AD.

**Conclusion:** EEG shows promise as a cost-effective, non-invasive tool for early detection and differential diagnosis. Future research should aim to collect standardized data from multicentric cohorts, across multiple diseases and stages, and explore the neural underpinnings of these diseases, to improve interpretability of the findings.

## Introduction

The estimated global prevalence of dementia in 2019, 57.4 million cases, is predicted to show a 166% increase by 2050 due to population growth and ageing [1]. Alzheimer’s Disease (AD) is the most common type, representing 60-70% of dementia cases, followed by Frontotemporal Dementia (FTD) and Lewy Body Dementia (DLB). One of the critical challenges in dementia, is an early and accurate diagnosis of neurodegenerative disease. Early detection is crucial for management of the disease and the effectiveness of disease-modifying therapies [2]. However, several issues complicate diagnostic accuracy. First, conventional structural diagnostic tools such as magnetic resonance imaging (MRI) and computer tomography (CT) are not sensitive to metabolic and functional alterations in the brain, occurring years before clinical onset ([3]). Second, traditional diagnostic tools are lacking in disease specificity, in particular when faced with syndromes high in clinical overlap. Last, cerebrospinal fluid (CSF) biomarkers and positron emission tomography (PET) scans, are valuable in providing insights in underlying pathology but are costly, invasive and not widely-accessible. As an alternative, electroencephalography (EEG) has emerged as a non-invasive, widely-accessible and cost-effective tool in the search for reliable biomarkers. EEG measures neuronal activity at high temporal resolution, allowing detection of early functional changes associated with ageing and neurodegeneration, delivering unique insights in the effects of neuropathology on neurophysiological mechanisms underpinning cognitive functioning.

In AD, the utility of resting state EEG (rsEEG) rhythms in wakefulness (eyes open or eyes closed) have been studied as candidate biomarkers, as they are non-invasive, cost-effective and do not require the performance of tasks, tackling problems with fatigue and motivation. Compared to healthy controls (HC), AD patients show slowing, i.e. the leftward shift in the power spectrum from higher (α, β, and γ) towards lower frequencies(δ and θ), as well as alterations in connectivity, complexity and synchronization of neural activity [4]. An alternative approach is task-based EEG, providing insights into the amplitude and latency of neural responses or event-related potentials (ERP) elicited by different cognitive tasks. A recent systematic review highlights reduced amplitude and delayed latency in AD compared to healthy controls in various well-known ERP components such as the P300 and N400 [5]. However, heterogeneity in the AD samples and study paradigms, along with scarcity of reported effect sizes, complicates meta-analyses across both resting-state EEG [4] as task-based EEG studies [5].Furthermore, diagnostic criteria have advanced from traditional neuropsychological and clinical measures to the inclusion of biomarkers, improving diagnostic accuracy in research studies and enhancing the reliability of research outcomes.

Another issue is the differential discrimination between neurodegenerative diseases. The differential diagnosis in early stages is suboptimal, for instance in discriminating DLB from AD. In the most recent consortium on diagnostic criteria, posterior slow-wave activity showing periodic fluctuations within the pre-α/θ range in resting-state EEG was recognized as a supportive biomarker for DLB [6]. From a recent systematic review, over 90% of DLB patients have diffuse EEG abnormalities [7]. Although EEG slowing was found to be more severe in AD compared to DLB, EEG abnormalities related to DLB overlap with those found in other neurodegenerative diseases such as AD. Just as in AD studies, previous systematic reviews in DLB have addressed issues such as preliminary sample sizes with large heterogeneity in patient cohorts, and lack of standardization in EEG protocols and reporting, complicating the identification of the optimal feature for differential diagnosis [7, 8]

In FTD research, studies have pursued similar aims of improving diagnosis with EEG, showing slowing of the power spectrum [9, 10]. However, the scarcity of studies in FTD results in too little evidence to reach consensus.

Although numerous studies have been conducted on the use of EEG in AD, FTD and DLB, no comprehensive review has yet compared EEG results across neurodegenerative diseases. Addressing this gap is crucial to define the specificity of electrophysiological markers in the differential diagnosis.

Additionally, to evaluate the utility of EEG in early stages of the diseases, we are interested in the preclinical and prodromal stages of the diseases. In line with the current diagnostic standards, we include exclusively biomarker-proven stages of AD [11]. To summarize the available literature, we conducted a systematic search on publications studying EEG-markers across AD, FTD, DLB and/or their preclinical and prodromal phases, compared to healthy controls.

## Methods

The review was prospectively registered in PROSPERO (ID: CRD42023392253). We performed a systematic search using online databases Pubmed, Cochrane, Web of Science and Scopus for English articles published from 2000 to January 2023. The full search strategy is provided in supplementary material (S1).

### Screening

Our search yielded a total of 12010 studies. The online collaboration platform Rayyan (https://www.rayyan.ai/) was used to implement, deduplicate and screen articles. Possible duplicates as detected by Rayyan were manually checked. Following deduplication, 6952 abstracts were screened for eligibility based on inclusion and exclusion criteria by two independent researchers (SDK, RB) blinded to each other’s rating. Based on abstract only, 6243 articles were excluded. The remaining 709 potential eligible articles were then screened on full text based on the same inclusion and exclusion criteria. The main reason of exclusion was not including biomarkers in AD (*n*=476). Conflicts were resolved by discussion between the researchers. The PRISMA flowchart (Figure 1) shows the full screening and exclusion process.

**Figure 1.**
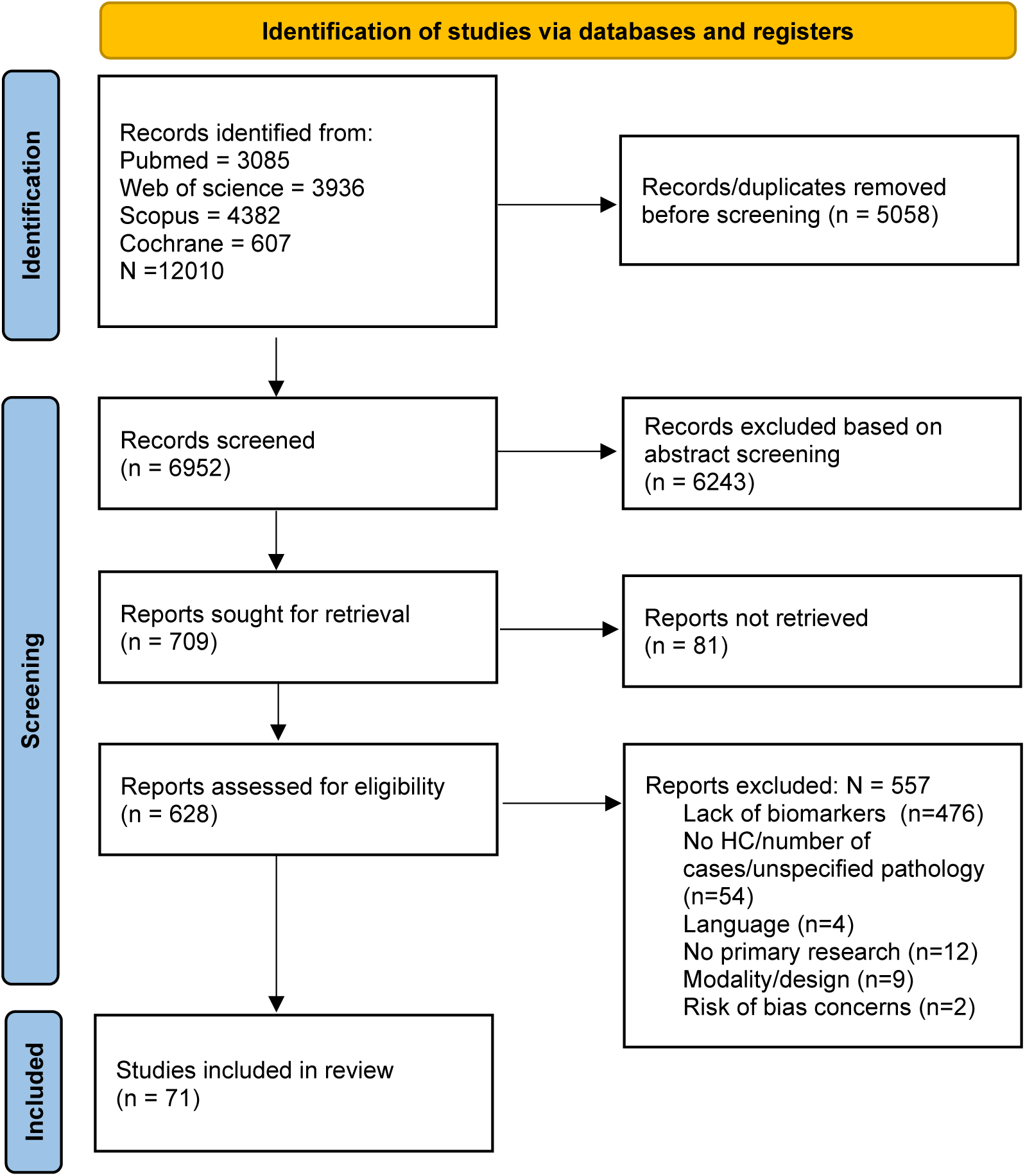
PRISMA flowchart diagram for systematic reviews [16].

#### Inclusion criteria

- Participant population: AD, FTD, DLB, including preclinical and prodromal stages, presymptomatic carriers of genetic causative mutations and healthy controls. In line with the International Working Group 2 (IWG-2) criteria for AD, we only include biomarker-proven cases of AD showing in vivo evidence of amyloid and tau pathology [12], presymptomatic carriers of familiar AD (FAD) by *Amyloid Precursor Protein (APP)* and *Presenilin (PSEN1/PSEN2)* causative mutations were included. Criteria for FTD diagnosis were a diagnosis following the Rascovsky criteria for bvFTD [13], Gorno-Tempini criteria for PPA[14], carrier of a genetic mutation of FTD, and/or imaging evidence of frontotemporal atrophy. DLB was defined by the criteria of McKeith [6, 15].

- Primary research studies

- English language

- Studies using resting-state EEG and/or task-based EEG as neuroimaging modality

#### Exclusion criteria

- Studies that combine multiple neuroimaging modalities or EEG with other biomarkers but do not separately report the performance of EEG markers.

- Animal studies

- Population with comorbidities (Down Syndrome, Schizophrenia, …)

- Prognostic studies

- Case studies or study population <5 participants

- Sleep EEG

Lastly, risk of bias was assessed using a hybrid version of the Joanna Briggs Institute (JBI) Critical Appraisal checklist (supplementary material S2).

### Data extraction

The following data was extracted manually from eligible studies. Tables with data can be found in supplementary material (S3).

1. Article information: First author, year
2. Study population: Sample size, patient population(s), control group(s), biomarkers
3. EEG acquisition: number of electrodes, sampling frequency, recording state (resting/task-based), duration
4. Analysis
5. Reported results

### Data synthesis

As our field of interest includes a heterogenous group of pathologies and disease stages as well as various recording and analysis conditions, data is presented descriptively. AUROC characteristics, sensitivity, specificity and diagnostic accuracy were reported if provided.

## Results

### Study characteristics

Our search yielded a total of 6957 unique studies, of which 71 studies were included in the final review (*figure 1*). In total, we included 34 AD studies, 9 FTD studies, 18 DLB studies, and 9 studies combining disease populations. The studies included a total of 1115 patients with dementia due to biomarker-proven Alzheimer (demAD) and 915 prodromal AD (prodAD) cases. Seven studies included preclinical AD pathology by positive amyloid and/or tau biomarkers (*n*=397). Six studies included FAD (*n*=43) and/or prodromal FAD (prodFAD, *n*=22) and asymptomatic carriers of a PSEN 1 or APP mutation (AcrFAD, *n*=100). Four studies included patients with bvFTD (*n*=108), three studies included not specified FTD (*n*=38). One study included familiar CHMP2B-FTD symptomatic (*n*=5) and presymptomatic carriers (*n*=5). One study compared non-fluent variant PPA (*n*=18), semantic variant PPA (*n*=10) and logopenic variant PPA (*n*=12). 18 studies included DLB patients (*n*=589), while only one study focused on prodDLB (*n*=21). In case studies included DLB or FTD combined with non-biomarker proven AD, only the results of the FTD or DLB patients are described. Five studies compared prodAD (*n*=111) and prodDLB (*n*=111), while one study included demAD (*n*=66) and DLB (n=66). Two studies using the same dataset compared PPA (*n*=5), bvFTD (*n*=13) and prodAD (*n*=18), a third study compared bvFTD (*n*=48) to AD (*n*=69). The majority of the studies employed a rsEEG recording (figure 2c). All of studies combining different disease populations employed resting state EEG.

**Figure 2.**
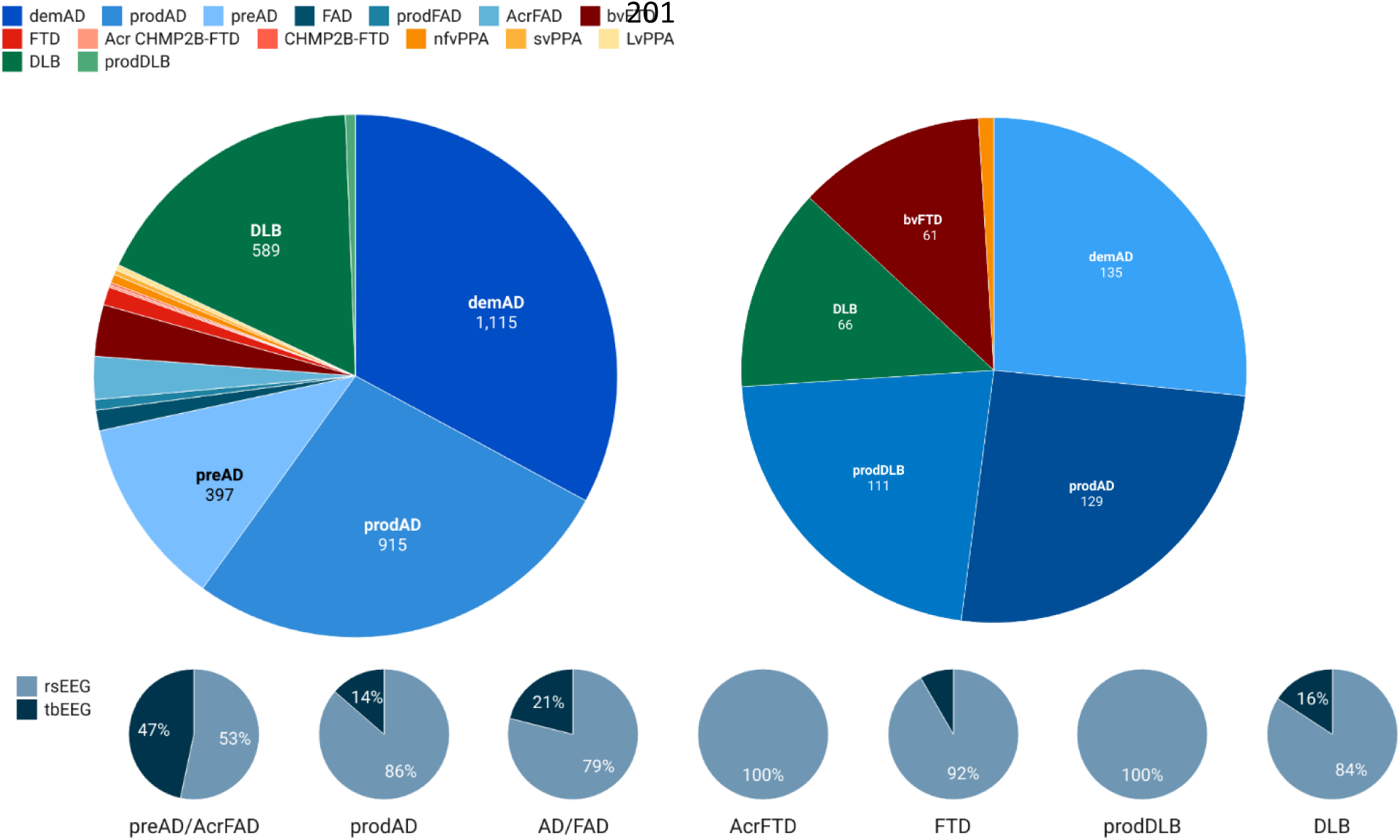
a) Study characteristics in studies including a single patient population. b) Study characteristics comparative studies. c) Percentage of studies using rsEEG or tbEEG per population. AD= Alzheimer’s Disease, prodAD= prodromal AD, preAD=preclinical ADFAD= Familial AD, prodFAD= prodromal FAD, ACR FAD= asymptomatic carriers of a FAD causative mutation, FTD= frontotemporal dementia, bvFTD= behavioral variant FTD, CHMP2B-FTD= symptomatic CHMP2B carriers, CHMP2B-FTD Acr= asymptomatic CHMP2B carriers, nfvPPA= non-fluent variant primary progressive aphasia (PPA), svPPA= semantic variant PPA, lvPPA= logopenic variant PPA, DLB= Dementia with Lewy Bodies, prodDLB= prodromal DLB, rsEEG= resting-state EEG, tbEEG= task-based EEG

### Potential for AD diagnosis

#### Resting state EEG

Diffuse slowing of the frequency spectrum towards lower frequencies is a consistent observation in both prodAD [17–20] and demAD [21, 22]. The shift in the power spectrum in demAD manifests as reduced spectral metrics in the α band, while spectral power in θ band increases [22]. Smailovic found that low α global field synchronization (GFS), a measure of global functional connectivity in the frequency domain, is linked to abnormal Aβ42 levels in demAD and prodAD, and high levels of p-tau and t-tau in prodAD [23]. Secondly, in demAD, high levels of p-tau and t-tau correlate with low global strength of scalp potential (GFP) in β and δ bands. In prodAD, high GFP in δ and θ was linked to abnormal CSF levels of Aβ42, while lower GFP α and β is linked to increased p-tau and t-tau. These results are in line with the research of Cecchetti, showing that prodAD show higher θ density than MCI without AD pathology with over 75% sensitivity and 70% specificity [21]. Similarly, Rodriguez demonstrated that frontotemporal alterations in β band discriminate between presymptomatic PSEN1 carriers and noncarriers and between FAD and HC (see figure 3)[24]. Additionally, significant progressive alterations in bispectral metrics are found along the AD continuum [25]. Specifically, with increasing severity, interactions between the δ and θ bands and other bands increase, while interactions with high frequency bands α, β1, and β2 bands diminish. Similarly, synchronization measures demonstrate reduced functional connectivity in demAD in α and β bands, while coherence metrics in θ band increase [26]. These results are similar to findings by Revilla-Vallejo, where Shannon Entropy (SE) shows higher values in the δ band and lower values in the α and β1 bands in prodAD and demAD compared to HC, suggesting less connectivity and integration in these two bands [27].

**Figure 3.**
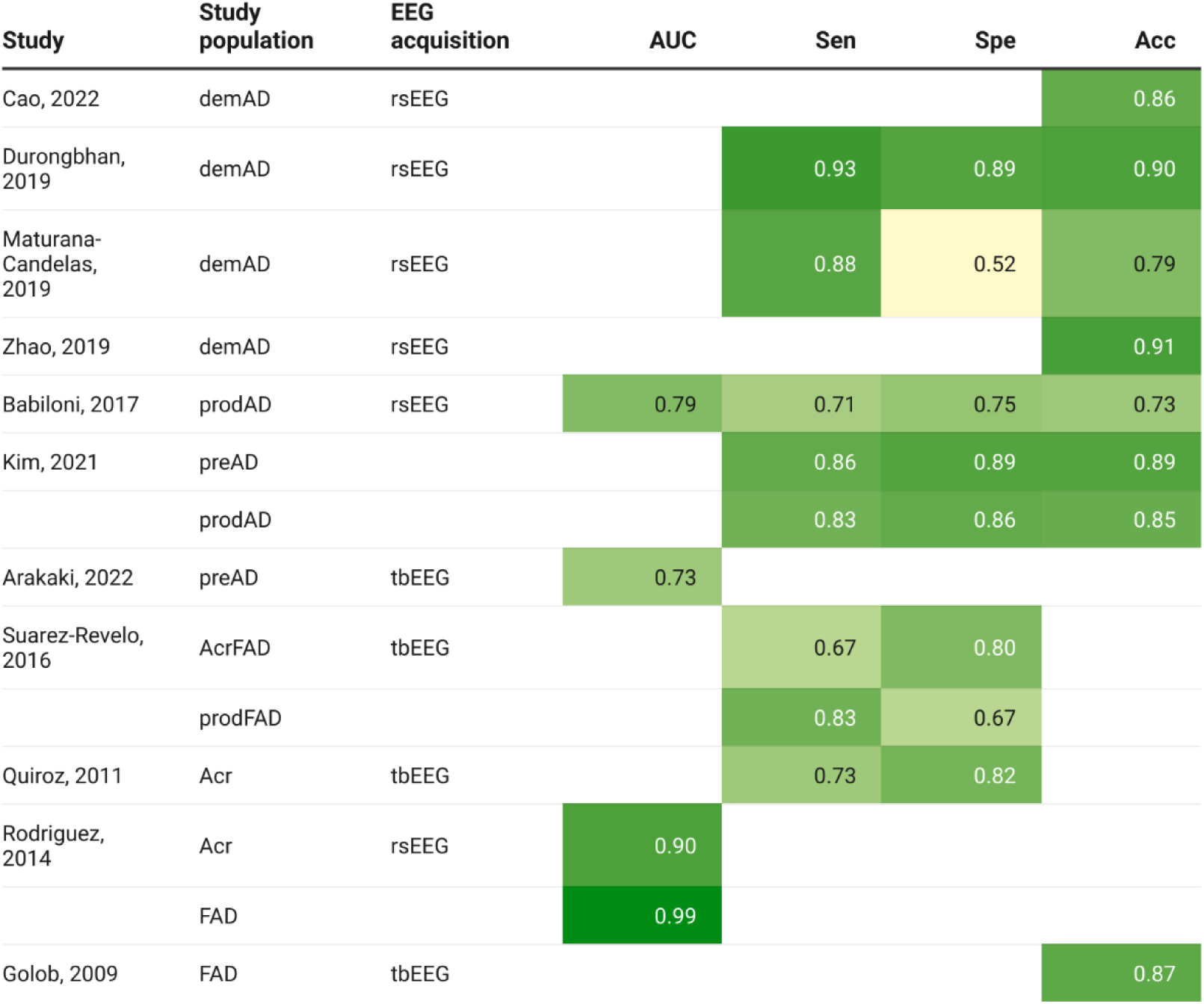
AUROC values in AD studies. If multiple values were provided, only the highest value was reported. Studies were organized according to subpopulation. AD=Alzheimer’s Disease, prodemAD= prodromal AD, preAD= preclinical AD, Acr = Asymptomatic carriers, prodFAD= prodromal Familial AD, FAD= Familial AD, rs-EEG= resting-state EEG, tbEEG= task- based EEG, AUC= Area under the curve, Sen=sensitivity, Spe= specificity, Acc= accuracy

Another way to look at resting-state activity is through microstate analysis. Spontaneous resting state activity can be described using microstates, transient global patterns of scalp potential, reflecting coordination of neural activity among networks [28]. Microstate analysis in AD reveals alterations in the duration, occurrence and coverage of microstates varying with severity. Specifically, demAD patients showed a longer duration in microstate B [29]. In the study of Smailovic, topographic differences in microstates A and D were found between HC and patient populations SCD, prodAD and demAD [30]. Furthermore, preclinical and prodromal AD could be differentiated from the dementia stage by topographical differences in microstate A. The authors further found that topographical alterations in microstate C were linked to increased Aβ42 levels, while p-tau levels were linked to microstate B alterations.

Recently, a number of studies used EEG-based machine learning classifiers for the detection of AD. An advantage of these studies is that the outcome measures offer more insight into the potential clinical implementation, which necessitates robust results at the individual level, compared to the prior studies which reported results from group-level comparisons. Studies discriminating AD from HC show promising classification accuracies in demAD ranging from 78% to 91% and in prodAD ranging from 73-85% (see figure 3). Another measure of the model’s accuracy is the F1 score, which is calculated using precision (positive predictive value) and recall (true positive rate). Studies achieved encouraging F1 scores ranging from 0.86 to 0.88 [31, 32]. Regarding the age factor, Durongbhan [33] found encouraging results in discriminating demAD from HC below the age of 70 (sensitivity >90%, specificity >83%), in individuals over 70 years, results were a little lower (>67% sensitivity, >85% specificity). Zhao studied both linear and nonlinear synchronization and found age- and disease-related differences in network synchronization [34]. Healthy individuals below 70 years old exhibit widespread linear synchronization with dynamic variability between eyes closed (EC) and eyes open (EO) states, while in young individuals with demAD, this dynamic variability is diminished, indicating network dysfunction. Secondly, they found a high widespread nonlinear synchronization during EO with higher dynamic variability compared to demAD. In individuals above 70, both HC and demAD showed similar levels of linear and nonlinear synchronization with minimal variability between states. Combining linear and nonlinear connectivity yields highest classification accuracies below the age of 70 (80.3% EO, 74.5% EC) and above the age of 70 (EO 86.5%, EC 90.5%).

In preAD, PSD patterns can be used to model compensatory mechanisms and progression to AD [35]. More specifically, individuals with amyloid burden in absence of neurodegeneration evidenced by brain metabolism in AD core regions, show increased functional connectivity in the parieto-occipital α band. In individuals with neurodegeneration, the impact on EEG metrics varies depending the degree of amyloid accumulation. More specifically, surpassing a critical threshold of amyloid accumulation, reverses the compensatory upregulation of higher β and γ frequencies and decreased δ power seen in intermediate amyloid burden, resulting in decreased β and γ power, MSF, spectral entropy, complexity and wSMI in θ band while δ power increases. In individuals with subjective complaints (SCD) with amyloid burden, Shim observed similar patterns of increased δ power in parietal, occipital and posterior cingulate regions combined with decreased α activity in fusiform and inferior temporal areas[36]. In predicting AD pathology in individuals with SCD, the best ML model shows 88.6% accuracy (see figure 3)[37].

#### Task-based EEG

A first important domain affected in AD is **encoding and memory**. Tautvydaite [38] shows that demAD patients show neural deficits in novelty detection and encoding during both learning and delayed memory recognition of pictures. Specifically, demAD patients show a decreased P200 response to new and repeated items, reflecting attention and perceptual processing, as well as an increased P300 during delayed repetition, which might reflect difficulties differentiating new from familiar stimuli. Similarly, in a passive picture recognition task of Stothart [39], demAD patients showed a decreased neural response to familiar pictures. Early differences in visual short term memory have also been found by Pietto[40] showing a reduced N1, P2 and P3 in prodFAD. Several studies including asymptomatic carriers of an AD causative genetic mutation, explored the potential of task-based EEG for early detection, highlighting early neural functional alterations without corresponding behavioral impairments. Golob [41] observed that presymptomatic carriers of the PSEN1 or APP mutation (*mean age* = 33.9), show discriminable neural alterations during an auditory target detection task. These alterations included decreased slow wave amplitudes, increased P200 amplitude, and delayed N100, N200, P200 and P300 latencies. Comparison of latencies across the ERP components showed that the latencies in the asymptomatic carriers were around 10% longer than the noncarrier group. Nontarget P200 latency emerged as a potent discriminator, successfully identifying 87% of the presymptomatic carriers. Quiroz [42] observed that during a recognition memory task, presymptomatic PSEN1 carriers showed lower frontal ERP positivity alongside an increased occipital positivity, with 72.7% sensitivity and 81.8% specificity discrimination. Despite these neural differences, both groups performed equally well on the task. Control subjects during recognition memory, exhibited activation patterns reliably associated with frontally mediated processes distinguishing between studied and unstudied visual items. PSEN1 carriers on the other hand, showed increased brain activity in occipital regions associated with visual perceptual processing. Ochoa [43] demonstrated a higher connectivity during the encoding condition of the same recognition memory paradigm in PSEN1 presymptomatic carriers compared to non-carriers. Connectivity in the occipito-parietal region during the same memory encoding condition within the 500-600ms time window is able to differentiate between presymptomatic and non-carriers with 67 % sensitivity and 80 % specificity [44]. P300 latency during an auditory oddball paradigm correlates with CSF levels of p-tau181, p-tau199 and ptau231 across demAD, prodAD and HC, while N200 latency negatively correlates with Aβ42 [45]. Recent studies in preAD using a working memory paradigm point towards reduced α event-related desynchronization (ERD) and altered α spectral entropy, suggesting compensatory hyperactivity during low load and insufficient cognitive resources with increasing work load [46, 47]. In the γ band, low working load induces a higher low γ in preAD, while decreases in γ are observed during high load [48].

In the **language domain**, sources of ERPs during a semantic-matching task reveal distinct topographical patterns in presymptomatic carriers versus non-carriers, although their behavioral responses and N400 amplitudes remain similar [49]. Specifically, presymptomatic carriers show a notable decrease in N400 generator strength within the right inferior-temporal and medial cingulate areas, and an increase within the left hippocampus and parahippocampus compared to non-carriers. The observed shift in N400 distribution mirrored that seen in symptomatic carriers, albeit with a less pronounced reduction in generator strength.

### Potential for FTD diagnosis

#### rsEEG

Spectral analysis in FTD shows a consistent slowing of the frequency spectrum. While studies agree on a slowing in α frequencies, there is some discrepancy regarding the changes in the β band, with some indicating a decrease and others suggesting an increase [9, 50]. In the study of Herzog [51], hypoconnectivity in the δ band between frontal, temporal, parietal and posterior areas emerged as the most relevant EEG feature, showing excellent AUC values in the discrimination of bvFTD from HC (figure 4). One study found right frontotemporal hypoconnectivity in bvFTD, which correlated with deficits in a naturalistic social text task [52].

**Figure 4.**
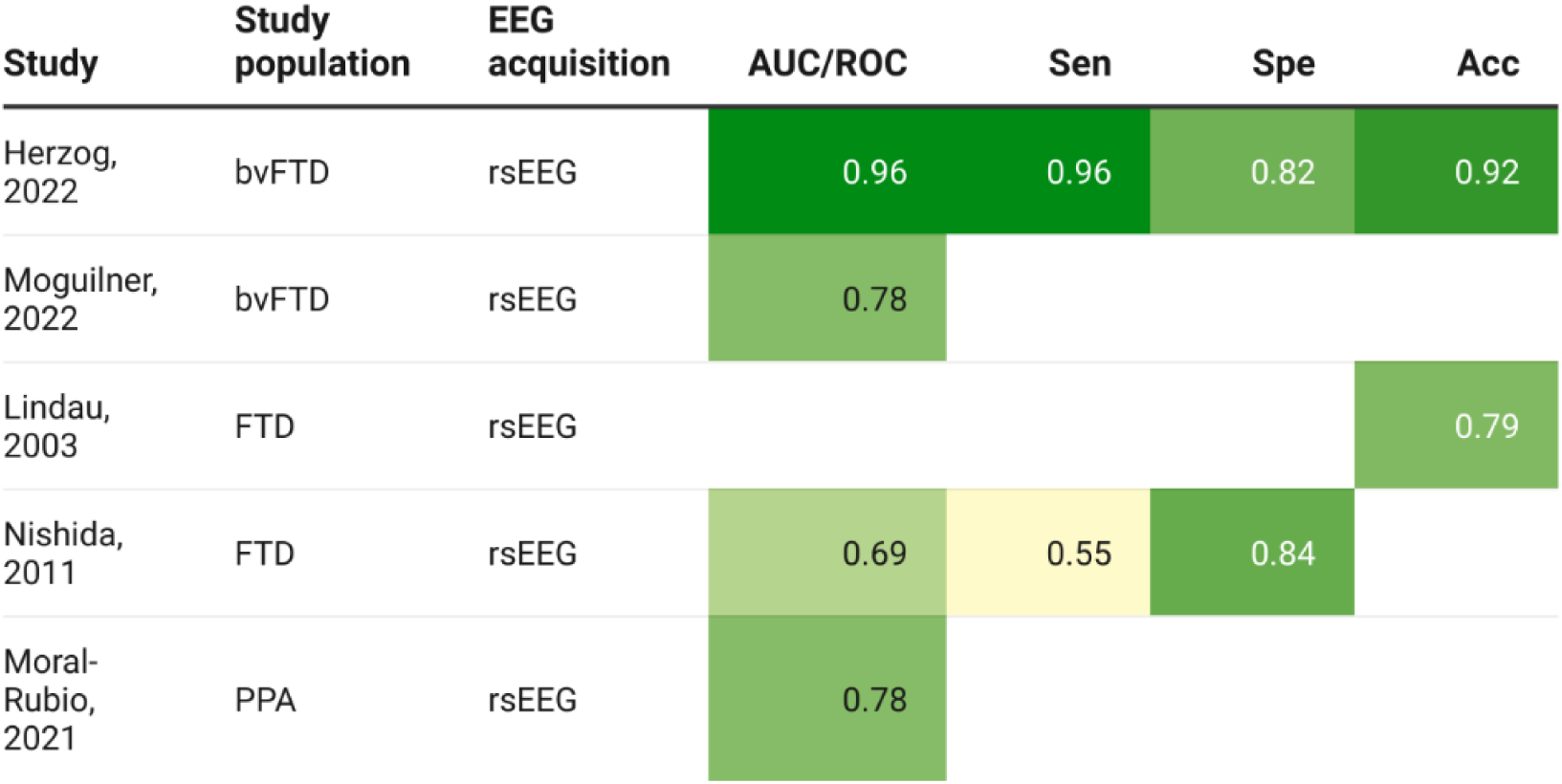
AUROC values in FTD studies. If multiple values were provided, only the highest value was reported. Studies were organized according to subpopulation. bvFTD= behavioral variant Frontotemporal Dementia, FTD= Frontotemporal Dementia, PPA= Primary Progressive Aphasia, rsEEG=resting-state EEG, AUC= Area under the curve, Sen=sensitivity, Spe= specificity, Acc= accuracy

Microstates were investigated in two studies. One study found that microstate duration C was decreased in FTD and that the sequence of activation from C to D was reversed [53]. Another study found that microstates vary with disease progression, showing an initial increase in microstate D, and a decrease as disease duration prolongs in CHMP2B-FTD [54]. These conflicting results could be explained by the different stages and variants of FTD. More specifically, microstate C is linked to the insular-cingulate network, linked to changes in personality typical of bvFTD. In contrast, CHMP2B-FTD presents with early impairments in executive functioning, with behavioural symptoms occurring at a later stage of the disease [54]. Microstate D has previously been linked to the fronto-parietal network, the initial increase followed by a decline in microstate D activation may be interpreted as a temporary compensatory mechanism. Studies using ML classification achieve encouraging AUC values between 0.78-0.96 for bvFTD and 0.78 for PPA versus HC (see figure 4). Moral-Rubio [55] also discriminated between nfvPPA, svPPA and lPPA variants, with 58% accuracy.

#### Task based EEG

Only one out of 10 studies included a task-based Go/No go paradigm, showing differences in θ and δ modulation related to impaired inhibition [56].

### Potential DLB

#### rsEEG

The most consistent finding in DLB compared to HC is slowing of the EEG pattern. More specifically, results show a leftward shift in mean dominant frequency (DF) from α range to pre-α or high-θ (6-7.5 Hz), in anterior [57], occipital [58, 59], posterior [60], or widespread areas [61]. Similarly, increased power in low frequency bands [61, 62] and decreased power in high frequency bands [61], as well as decreased θ/α ratio [58], differences α/θ and α/δ ratio [60] are observed. Using the θ/α ratio, DLB patients can be discriminated from HC with 76.7% sensitivity, 61.7% specificity and 66.7% accuracy [63]. In DLB patients with visual hallucinations, parietal δ activity achieves 75% sensitivity, 85% specificity, 81% accuracy in discrimination from HC. Posterior α reactivity from EC to EO is reduced in DLB, suggesting altered neural vigilance in the occipital lobe, enabling discrimination of DLB with 95.2% sensitivity [64].

In terms of connectivity, DLB patients show consistent network disorganization. DLB was characterized by network randomization and reduced connectivity in the α and β band [59, 65, 66] combined with increased network complexity in the high-θ band [59]. Similarly, combining connectivity strength of the β band with graph features of α band achieved an 76% accuracy in differentiating DLB from HC in the study of Mehraram [67]. In terms of dynamic connectivity, DLB patients show reduced α band information flow between posterior to anterior areas [68]. Functional source connectivity suggests cortical disconnection in DLB as both intra- and interhemispheric lagged linear connectivity (LCC) in the α range are reduced. Interhemispheric LCC in α range achieves good discrimination [69]. In the study of Kai [62], decreases in interhemispheric coherence (Icoh) and intrahemispheric coherence (Hcoh) were besides α also pronounced for δ, β and θ frequencies. In terms of large-scale resting state networks (RSN), connectivity decreases are found in the visual and sensorimotor network [65].

Two studies on microstates found contrasting results. In the study of Schumacher [70] microstate duration of all microstates was increased with reductions in numbers of microstates per second in DLB. Lamos [71] found the opposite direction of changes in prodDLB, with increased occurrence of all microstates, and shorter mean duration and increased occurrence of microstate B, which is associated with the visual network.

#### Task-based EEG

Three studies included a task-based EEG paradigm. One study used an auditory oddball paradigm and found a reduced and delayed P300 amplitude, as well as a P300 latency gradient inversion in DLB [72]. A longer latency of the P300 was also found in a visual oddball task [73]. Interestingly, oddball tasks reveal differences in EEG oscillations reflecting typical DLB symptoms. More specifically, DLB patients show a decreased event-related δ synchronization combined with impaired α and β suppression during both visual and auditory oddball tasks, and decreased θ band activity in a visual task [73, 74]. From a clinical perspective, decreased event-related θ oscillations and decreased α suppression during a visual oddball task may serve as neurophysiological correlates of attention and visual dysfunction in DLB. Power in δ band was able to discriminate DLB from HC with good sensitivity and specificity [74].

### Potential for discrimination

#### AD versus DLB

Studies comparing prodAD and prodDLB consistently find a more pronounced slowing in prodDLB compared to prodAD [75–77]. To account for interindividual variability, individual α frequency peaks (IAF, maximum power peak between 6-14Hz) and transition frequency between α and θ band (TF, minimum power density between 3-8Hz) can be used. Reduced mean TF and mean IAF values are consistent observations across DLB studies [63, 69, 70]. In the study of Babiloni [78] mean TF and mean IAF were found to be lowered in both prodAD (*m*IAF=8.8Hz, *m*TF = 5.4) and prodDLB compared to HC (*m*IAF = 9.4Hz, *m*TF=96.3Hz) with more pronounced reductions in prodDLB (*m*IAF =7.8Hz, *m*TF =4.7Hz). In the discrimination of prodAD from prodDLB, sensitivity values for spectral metrics range between 41 to 78.3%, while specificity values range between 66.7 to 97% (see figure 6) [76, 78]. The leftward shift in the power spectrum in DLB was also confirmed in the study of Massa [75], where α/θ ratio was found to be decreased in prodDLB compared to prodAD, and in the study of Schumacher [76], where increases in θ/α ratio were correlated with more severe cases of prodDLB. In the study of Schumacher [76], β power showed the highest AUC (0.71) in discriminating prodAD from prodDLB, with 61% sensitivity and 81% specificity. Discrimination between DLB and AD with high β power as the most important factor, shows good sensitivity and specificity (see figure 6) [68]. Babiloni [79] found that inter- and intrahemispheric LCC values in the α band were reduced in both prodAD and prodDLB compared to HC. While intra-hemispheric LCC α2 was best to differentiate prodDLB from HC, interhemispheric LCC global α2 was best to differentiate prodAD from HC. However, discrimination between prodAD and prodDLB remained low (AUC <0.7).

#### AD versus FTD

While both FTD and AD show progressive connectivity alterations compared to HC, profound frequency- and location dependent differences can be found in network organization, which are linked to the core areas of the diseases. Compared to bvFTD, demAD patients show lower connectivity in the α and δ band in posterior regions and a widespread higher connectivity in the θ band [80]. bvFTD patients showed an inverse pattern, with preserved posterior alpha connectivity, but lower θ activity in posterior and frontal areas. The Minimum Spanning Tree (MST) analyses indicate that frontal networks appear to be selectively involved in bvFTD, whereas in AD, global efficiency is reduced [80]. In the study of Franctiotti [81], the typical main hub in HC is lost in FTD at dementia onset and replaced by frontal local hubs, while network organization is largely preserved in demAD. In the same study, global clustering was able to distinguish between FTD (PPA and bvFTD) and demAD with moderate sensitivity but high specificity (see figure 6). In the study of Bonanni [82], network alterations predominantly targeted the frontal region in pFTD, while in prodAD, mutual information in the left local anterior region discriminated prodFTD from prodAD with good sensitivity but low specificity (see figure 6). Using the same metric, discrimination of pFTD from HC reached 89% sensitivity and 90% specificity. For the discrimination of prodAD from HC, MI in the posterior connections achieved high sensitivity (89-100%) and specificity (85-100%). Interestingly, these effects were evident in the prodromal stages of the disease but diminished with progression, which is suggestive of hyperconnectivity as a temporary compensatory mechanism to account for the effects of neurodegeneration in the core areas of the specific disease.

**Figure 5.**
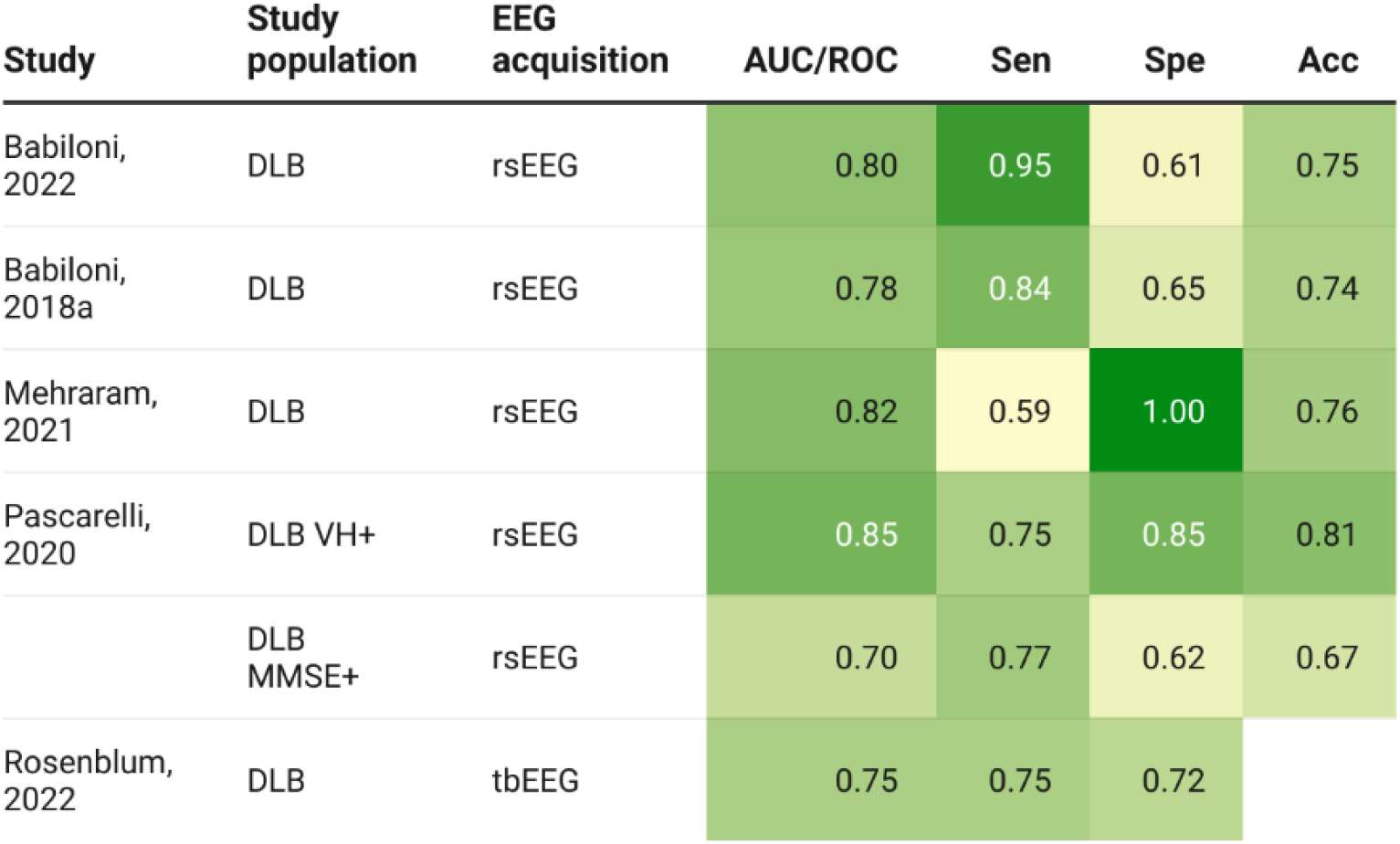
AUROC values in DLB studies. If multiple values were provided, only the highest value was reported. Studies were alphabetically organized. DLB= Dementia with Lewy Bodies, DLB VH+= DLB with visual hallucinations, DLB MMSE+= DLB with pathological Mini Mental State Examination scores, rsEEG= resting-state EEG, tbEEG= task-based EEG, AUC= Area under the curve, Sen=sensitivity, Spe= specificity, Acc= accuracy.

**Figure 6.**
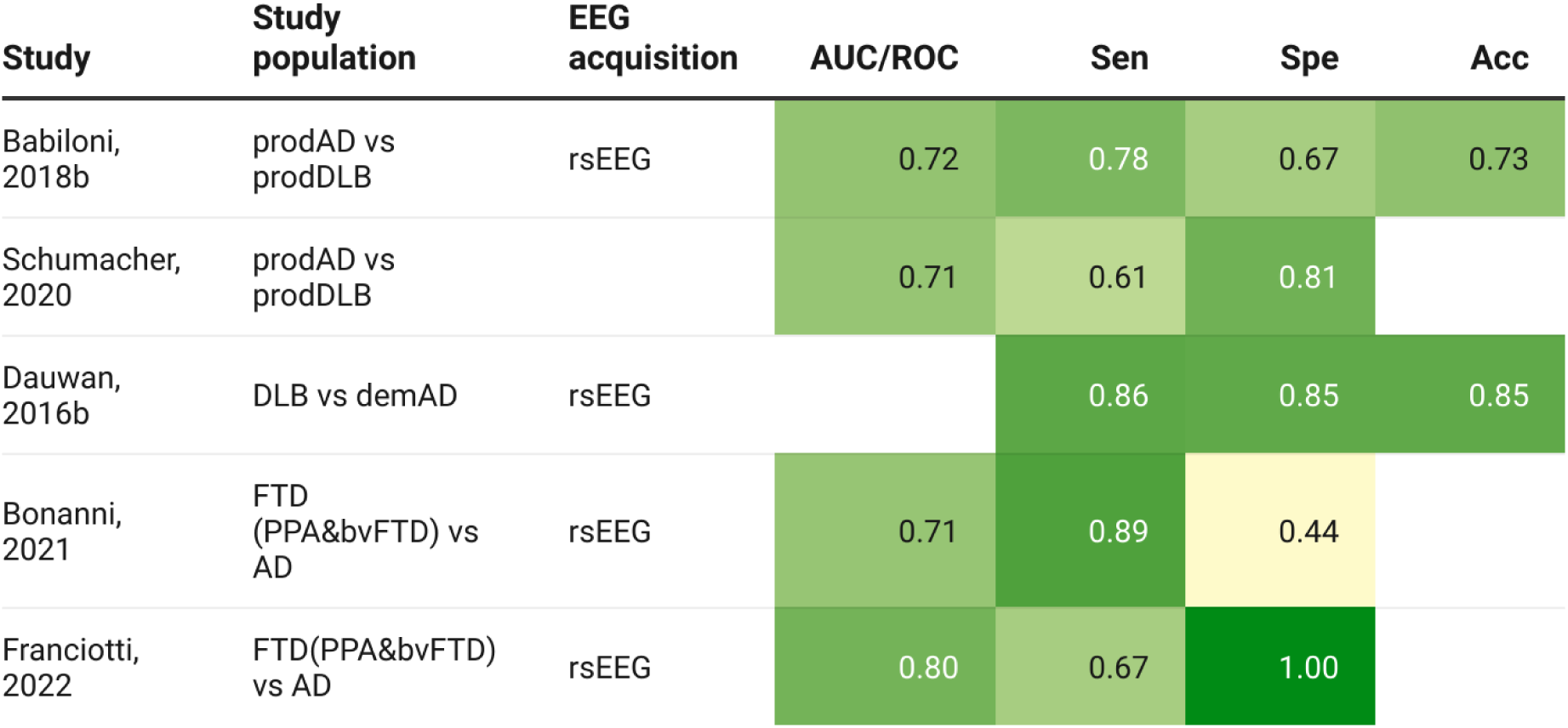
AUROC values in discriminative studies. If multiple values were provided, only the highest value was reported. prodAD= prodromal Alzheimer’s Disease, prodDLB= prodromal Dementia with Lewy Bodies, prodFTD= prodromal Frontotemporal Dementia, DLB= Dementia with Lewy Bodies, demAD= dementia due to Alzheimer’s Disease, PPA= Primary Progressive Aphasia, bvFTD= behavioral variant Frontotemporal Dementia, rsEEG= resting-state EEG, tb-EEG=task-based EEG, AUC= Area under the curve, Sen=sensitivity, Spe= specificity, Acc= accuracy.

## Discussion

To the best of our knowledge, this is the first systematic review to evaluate the diagnostic utility of EEG across AD, FTD, DLB as well as their preclinical and prodromal stages. The findings reveal promising AUC values across diseases and disease stages, which suggest that EEG holds significant diagnostic utility in AD, FTD and DLB. This, in combination with the advantages of EEG such as cost-effectiveness and non-invasiveness, could make EEG a valuable tool for the detection of neurodegenerative diseases.

### Potential for diagnosis of AD, FTD and DLB at the dementia stage

In AD, the leftward shift towards lower frequencies is a well-established finding, characterized by reductions in spectral and connectivity metrics in higher frequencies α and β, while increases in these metrics are found in δ and θ bands [21–23, 25–27]. Discrimination of AD based on frequency data is promising with excellent sensitivity values ranging from 88 to 93%, albeit lower specificity between 52-89% (see figure 3). In DLB, similar patterns of slowing arise as the most consistent findings [57–63, 69, 70]. Discrimination of DLB from HC using spectral metrics ranges between 75-95% sensitivity, 61-85% specificity and 66.7-81% accuracy (see figure 5). Similarly, reduced connectivity is found, especially within the α range [59, 62, 65–69]. Depending on the analysis, discrimination based on connectivity measures is lower with sensitivity values of 59-84%, specificity of 65-100%, and accuracy of 74-76% (see figure 5). Overall, the high sensitivity values suggest that frequency data holds promise as a screening tool in both AD and DLB.

Preliminary studies on FTD (bvFTD and PPA) have reported varying AUC values (ranging between 0.69-0.96, figure 4), however, due to the heterogeneity of the FTD spectrum, the variation in analysis techniques and the limited amount of studies, direct comparisons of AUC values are challenging. In bvFTD, connectivity analysis achieves the most encouraging AUC values [51, 83]. A different promising avenue could be microstate analysis, with alterations in microstate C and D, varying according to the subtype and stage of the disease [53, 54]. However, without ROC analysis to validate its effectiveness, the value of microstate analysis remains uncertain.

### Potential for early diagnosis

In prodAD, slowing of the frequency spectrum is the most consistent observation with slowing patterns similar to those observed in AD, with the most prominent and consistent reductions in alpha, TF and IAF [17, 18, 20, 21, 23]. Discrimination of prodAD from MCI without AD biomarkers and HC achieves moderate sensitivity and specificity values (see figure 3). One explanation might be that alterations in spectral metrics start before the start of preclinical AD and develop differently across individuals. Recent studies have reported interindividual variability in EEG metrics, depending on the degree of amyloid burden and neurodegeneration [35] as well as the interaction with other factors modulating brain activity such as age [19] and education levels [18, 84].

Research on early stages of FTD is limited, with only one study including presymptomatic carriers of a FTD causative mutation, suggesting progressive alterations in microstates [54]. More precise, the upregulation of microstate D activity in the early stages of the disease might be a temporal compensatory mechanism. However, as the disease progresses and the brain’s capacity to maintain this compensation diminishes, microstate D activity decreases. Similarly, microstate alterations have been suggested as a potential early marker for prodDLB, with alterations inverse to those in the dementia stage of the disease, which again may be explained by early maladaptive mechanisms [71].

### Potential for differential diagnosis

Another key issue is the differential diagnosis of neurodegenerative diseases which is complicated due to overlap in clinical symptoms and heterogeneity of the diseases. A leftward shift in the power spectrum is a consistent finding in all three populations, AD [17–22], FTD [9, 50] and DLB [57–62], raising questions regarding the specificity of this phenomenon. Studies comparing prodAD and prodDLB consistently find a more pronounced slowing in prodDLB, with good specificity values (66.7-97%) for the alpha band but low sensitivity (41-78.3%). In the study of Schumacher (2020), sensitivity values range between 23 and 61%, while specificity were higher between 81 and 89% for different frequency bands. The high specificity across studies suggests that a significant shift in the power spectrum is more likely to be indicative for prodDLB rather than prodAD. However, the low sensitivity values, indicate that in cases with less disturbed EEG patterns, differentiating between the two becomes increasingly difficult. As the disease progresses, discrimination between DLB and AD shows better sensitivity (86%) and specificity (85%) [85]. Interestingly, alterations link to the key areas of the diseases. In DLB, the slowing of EEG frequencies is more pronounced in posterior regions [58–60]. In FTD, reductions in alpha band are pronounced in frontal areas, which corresponds to anatomical and metabolic changes in these areas in FTD patients [50]. Connectivity studies provide similar evidence. In FTD, network alterations predominantly target frontal regions [80–82], aligning with the progressive frontal disconnection characteristic of the disease. In contrast, more widespread or posterior alterations seem to be more suggestive of AD [80, 82]. In DLB, functional connectivity is reduced in the visual and sensorimotor network [65]and information flow is reduced between posterior to anterior regions [68].

### Potential of task-based versus resting state EEG

While resting-state EEG measures spontaneous activity, task-based EEG captures the complex dynamics underlying cognitive processes that are affected in neurodegenerative diseases, thereby increasing interpretability. Task-based paradigms offer unique insights into the neural substrates of AD-related symptoms, showing alterations in auditory and visual target detection and memory recognition [38, 39, 41, 45]. Notably, task-based EEG has been studied extensively in the earliest stages of AD (figure 1c). Task-based EEG highlights early, subtle changes in neural processing of memory and language preceding deficits in performance in presymptomatic FAD carriers [41–43, 49]. More specifically, early alterations are present in well-known ERP components N100, N200, P200 and P300 that are also found in prodFAD [40] and demAD [38]. Sensitivity values of task-based paradigms identifying carriers from noncarriers range from 67-72.7% and 80-81.8% specificity (see figure 3), which is comparable to symptomatic stages. The ability to detect early neural changes before the onset of symptoms is perhaps one of the most promising findings, as it could revolutionize how we approach AD.

Three studies in DLB employed task-basedEEG, offering insights into the neural substrates of cognitive deficits in DLB. During oddball tasks, reduced and delayed P300 amplitudes are found, relating to deficits in attention and executive functioning [72, 73]. Furthermore, DLB patients show a decreased event-related δ and θ synchronization combined with impaired α and β suppression, which may serve as a neurophysiological correlate of visual and attentional dysfunction in DLB showing promising sensitivity and specificity values [73, 74]. We found no task-based studies discriminating between neurodegenerative diseases, despite evidence of alterations in task-based analyses, such as delayed P300 effects which have been reported in both AD [41, 45] and DLB [72, 73].

## Limitations

The available academic literature on the diagnostic utility of EEG markers between AD, FTD and DLB shows notable gaps. First, there is an underrepresentation of studies including FTD patients, which limits the ability to make statements about the most promising avenue in diagnosing FTD and discriminating FTD from other neurodegenerative diseases. Furthermore, there is a scarcity of studies comparing across neurodegenerative diseases, which limits the ability to make statements on the specificity of some markers.

The established heterogeneity within the AD, FTD and DLB population is thought to play a major role in the sometimes conflicting outcomes of prior research. As is shown in our review, not all disease stages within these neurodegenerative diseases are equally represented. Studies including multiple disease stages have shown differences in the EEG markers with progression of the disease. As not all studies describe disease duration, conflicting results might be partially explained by different disease stages. On the other hand, it is clear that other factors which are hard to model in their entirety impact disease presentation and severity besides disease duration. In our bias assessment, we paid special attention to the matching of confounding factors such as age and education that were studied in multiple studies. Education has been shown to influence alpha activity, serving a neuroprotective and compensatory role in AD [84]. A parallel pattern can be found between brain activity and cognitive reserve (CR), the brain’s resilience to neuropathology and neurodegeneration in prodAD [18]. In studies using a working memory paradigm, CR translated into better neural efficiency, evidenced by increased α ERD and decreased α SpecEn during high working load [47]. Taken together, these findings suggest both a compensatory mechanism in mitigating consequences of brain slowing in AD. Finally, the impact of neuropathological co-pathology remains largely uncharted territory.

In terms of methodology, the variety in markers of interest, methods and analysis along with a scarcity in reporting of AUROC values, severely limits the possibility for meta-analysis and statements about the most promising marker. Recent work shows promise in harmonizing EEG data across centers using novel post-processing methods [86].

The interpretation of EEG markers and their link to clinical symptoms remains largely unclear. Research exploring the correlation between neural mechanisms and clinical symptoms is particularly valuable in increasing the interpretability of neural alterations. For example, spectral and connectivity measures may clarify the association between DLB pathology and clinical symptoms. Multiple authors have proposed that a cholinergic deficit underlies α band network alterations [59, 65, 66]. Since α oscillations are involved in thalamo- and corticocortical communication crucial for cognitive processing, it is speculated that α band network alterations may connect cholinergic deficits to key clinical symptoms in attention, perception and memory [59, 65, 66]. Finally, task-based EEG shows a clear advantage to mechanistically study specific cognitive processes at the earliest disease stages, when neural changes are still relatively limited.

## Conclusions and future directions

In conclusion, EEG markers show promising AUC values for detecting AD, FTD and DLB. For the detection of early neural changes, task-based EEG markers are particularly valuable in identifying the earliest stages of AD, revealing neurophysiological changes before clinical symptoms become apparent. While promising sensitivity values are reported in the diagnosis of AD, FTD and DLB, the specificity of these biomarkers ask for further research. The most established marker, slowing of the EEG spectrum, seems to be rather a marker of neurodegeneration than specific to a certain disease.

However, frequency-dependent connectivity aids interpretation, illustrating disease-specific alterations corresponding with the core regions and symptoms affected by each disease. Exploration of both spectral metrics and microstates could be beneficial, especially with the intent towards discrimination from early stages of AD, which is important for clinical application.

To reduce heterogeneity in sample sizes, future studies should strive for clear descriptions of disease duration and demographic factors such as age and education levels. There is a growing body of evidence illustrating the influence of interindividual variability on the onset and progression of AD. Given the complex nature of disease progression, the modulatory and compensatory mechanisms of these diseases should be further explored. Furthermore, future research must validate previous findings in multicentric studies combining disease populations. Equally important will be the standardization of research protocols, including up-to date consensus criteria, and reporting of methods allowing replication and enhancing generalizability and interpretability.

In conclusion, the use of EEG shows promise in diagnostic accuracy and differential diagnosis and shows advantages in non-invasiveness, availability and cost-effectiveness. With further research, the search for the most optimal EEG marker could revolutionize the diagnosis of AD, FTD and DLB, establishing an early and accurate detection of neurodegeneration.

## Data Availability

All data produced in the present work are contained in the manuscript

## Acknowledgements

SVM, TVDB, DC, PC and RB are members of the European Reference Network for Rare Neurological Diseases - Project ID No 101085584.

## List of abbreviations

Ab42: Amyloid plasma
AEC: amplitude envelope correlation
AEC-c: AEC with leakage correction
AMI: Auto mutual information
ApEn: Approximate entropy
BispEn: bispectrum cubic entropy
BispMF: bispectrum median frequency
BispRP: Bispectrum relative power
CC: Clustering Coeficient
Coh: Mean global coherence
Cross-ApEn: Cross-Approximate Entropy
CSA: Compressed spectral arrays
CSD: Current Source Density
D2: Mahalanobis distance
DAR: δ to α ratio
DF: Dominant frequency
DFV: Dominant frequency variance
DRC: Dynamic Range of Connectivity
DT: Decision Tree’s
DTF: directed transfer function
DWT: Discrete Wavelet Transform
EA: EEG abnormalities
ERD: Event related desynchronization
ERO: Event Related Oscillations
ERS: Event Related synchronisation
FC: Functional Connectivity
FP: Frequency Prevalence
GEV: global explained variance
GFP: Global Field Power
GMFP: Global Mean Field Power
GSA: Gready search algorithm
GTA: Graph Theory Analysis
HC: Hjorth Complexity
HHT: Hilbert Huant transform
HOFC: Higher order functional connectivity
IAC: instantaneous amplitude correlation
iCoh: Imaginary coherence
ITPC: inter-trial phase clustering
kNN: k-nearest neighbour
LDA: Linear discriminant analysis
LLC: Lagged linear coherence
LOOcv: Leave one out cross validation
LOSO: Leave one subject out (cross-validation)
LR: Logistic Regression
LZC: Lempel-Ziv Complexity
MF: Median frequency
MI: Mutual information
MSCOH: magnitude squared coherence
MSE: Multiscale Sample Entropy
MSSE: Multiscale SpecEn
MST: Minimum Spanning Tree
MWC: Morlet wavelet convolution
NB: Naive Bayes
NCA: neighbourhood component analysis
NDTF: non-normalized directed transfer function
PCA: Principal Component Analysis
PL: Path Length
PLI: Phase Lag index
PLV: Phase Locking Value
PSA: Power Spectrum analysis
PSD: Power Spectrum Density
PSI: Phase synchronization index
PTE: Phase Transfer Entropy
QDA: Quadratic Dicriminant Analysis
RCG: Revised Circular Graph
RF: Random Forest
RMS: Root mean square
rMSSE: refined MSSE
RNN: recurrent neural network
RP: Relative Power
RSN: Resting State Network
SampEn: Sample Entropy
SE: Shannon Entropy
SMR: Stepwise multilinear regression
SNR: Signal to noise ratio
SP: Spectral power
SpecEn: Spectral Entropy
SPR: Statistical Pattern Recognition
SR: Spectral Ratio
SVM: Support Vector Machine
TAR: Theta to alpha ratio
TAS: temporal activation sequence (TAS)
TBR: Theta to beta ratio
TF: Transition Frequency
wPLI: weighted PLI
wSMI: weighted symbolic mutual information
WT: Wavelet Transform

