## Supplementary material for "Diagnostic Utility of Electrophysiological Markers for Early and Differential Diagnosis of Alzheimer’s, Frontotemporal, and Lewy Body Dementias: A Systematic Review"

**S1**

***Pubmed***

((("Electroencephalography"[Mesh] OR "Evoked Potentials"[Mesh] OR "EEG"[Title/Abstract] OR "Electroencephalogr*"[Title/Abstract] OR "Evoked potential*"[Title/Abstract] OR "Event Related Potential*"[Title/Abstract] OR "Event-Related Potential*"[Title/Abstract] OR "ERP"[Title/Abstract] OR "N1"[Title/Abstract] OR "N300"[Title/Abstract] OR "P2 wave"[Title/Abstract] OR "N100"[Title/Abstract] OR "N3"[Title/Abstract] OR "N400"[Title/Abstract] OR "N4"[Title/Abstract] OR "P2"[Title/Abstract] OR "P600"[Title/Abstract] OR "N2"[Title/Abstract] OR "N200"[Title/Abstract])

AND

("Lewy Bodies"[Mesh] OR "Lewy Body Disease"[Mesh] OR "Alzheimer Disease"[Mesh] OR "Frontotemporal Lobar Degeneration"[Mesh] OR "Frontotemporal dementia*"[Title/Abstract] OR "frontotemporal lobar degeneration"[Title/Abstract] OR "frontotemporal lobe dementia*"[Title/Abstract] OR "Semantic Dementia*"[Title/Abstract] OR "FTD"[Title/Abstract] OR "FTLD*"[Title/Abstract] OR "Alzheimer*"[Title/Abstract] OR "senile dementia*"[Title/Abstract] OR "presenile dementia*"[Title/Abstract] OR "Mild Cognitive Impairment"[Title/Abstract] OR "MCI"[Title/Abstract] OR "LBD"[Title/Abstract] OR "DLB"[Title/Abstract] OR "Lewy bod*"[Title/Abstract])

AND (2000:2023[pdat])) NOT (review[Publication Type])) NOT (systematic review[Publication Type])

***Cochrane (via Ovid)***

***Register of Controlled Trials***

(exp Electroencephalography/ OR exp evoked potentials/ OR (eeg OR Electroencephalogr* OR "Evoked potential*" OR "Event Related Potential*" OR "Event-Related Potential*" OR ERP OR N1 OR N300 OR "P2 wave" OR N100 OR N3 OR N400 OR N4 OR P2 OR P600 OR N2 OR N200).ti,ab,kw)

AND

(Lewy Bodies/ OR Lewy Body Disease/ OR Alzheimer Disease/ OR exp Frontotemporal Lobar Degeneration/ OR ("Frontotemporal dementia*" OR "frontotemporal lobar degeneration" OR "frontotemporal lobe dementia*" OR "Semantic Dementia*" OR FTD OR "FTLD" OR "Alzheimer*" OR "senile dementia*" OR "presenile dementia*" OR LBD OR DLB OR " Lewy bod*" OR “mild cognitive impairment”).ti,ab,kw)

limit 1 to yr="2000 - 2023"

Web of Science

(TS=((EEG OR Electroencephalogr* OR "Evoked potential*" OR "Event Related Potential*" OR "Event-Related Potential*" OR ERP OR N1 OR N300 OR "P2 wave" OR N100 OR N3 OR N400 OR N4 OR P2 OR P600 OR N2 OR N200)

AND

("Frontotemporal Dementia*" OR "frontotemporal lobar degeneration" OR "frontotemporal lobe dementia*" OR "Semantic Dementia*" OR FTD OR FTLD OR Alzheimer* OR “Mild Cognitive Impairment" OR MCI OR "senile dementia*" OR "presenile dementia*" OR LBD OR DLB OR "Lewy bod*"))) NOT DT=(Review) AND PY=(2000-2023)

Scopus

TITLE-ABS-KEY ( eeg  OR  electroencephalogr*  OR  "evoked potential*"  OR  "event related potential*"  OR  "event-related potential*"  OR  erp  OR  n1  OR  n300  OR  "p2 wave"  OR  n100  OR  n3  OR  n400  OR  n4  OR  p2  OR  p600  OR  n2  OR  n200 )  AND  TITLE-ABS-KEY ( "frontotemporal dementia*"  OR  "frontotemporal lobar degeneration"  OR  "frontotemporal lobe dementia*"  OR  "semantic dementia*"  OR  ftd  OR  ftld  OR  alzheimer*  OR “mild cognitive impairment" OR mci OR  "senile dementia*"  OR  "presenile dementia*"  OR  lbd  OR  dlb  OR  "lewy bod*" )  AND  PUBYEAR  >  1999 AND  ( LIMIT-TO ( DOCTYPE ,  "ar" ) )

**S2**

|  |  | **Were the criteria for inclusion in the sample clearly defined?** (yes, no, unclear, not applicable) | **Were objective, standard criteria used for measurement of the condition** (yes, no, unclear, not applicable) | **Were the study subjects and the setting described in detail?** (yes, no, unclear, not applicable) | **Were the groups comparable other than the presence of disease?** (yes, no, unclear, not applicable) | **Was there clear reporting of the demographics of the participants in the study?** (yes, no, unclear, not applicable) | **Were the outcomes measured in a valid and reliable way?** (yes, no, unclear, not applicable) | **Was appropriate statistical analysis used?** (yes, no, unclear, not applicable) | **Overall appraisal based on risk of bias (include/exclude)** |
| --- | --- | --- | --- | --- | --- | --- | --- | --- | --- |
| Aoki | 2019 | Yes | Yes | Yes | Yes | Yes | Yes | Yes | Include |
| Arakaki | 2019 | Yes | Yes | Yes | Yes | Yes | Yes | Yes | Include |
| Arakaki | 2022 | Yes | Yes | Yes | Yes | Yes | Yes | Yes | Include |
| Babiloni | 2017 | Yes | Yes | Yes | Yes | Yes | Yes | Yes | Include |
| Babiloni | 2018a | Yes | Yes | Yes | Yes | Yes | Yes | Yes | Include |
| Babiloni | 2018b | Yes | Yes | Yes | Yes | Yes | Yes | Yes | Include |
| Babiloni | 2019 | Yes | Yes | Yes | Yes | Yes | Yes | Yes | Include |
| Babiloni | 2020 | Yes | Yes | Yes | Yes | Yes | Yes | Yes | Include |
| Babiloni | 2021a | Yes | Yes | Yes | Yes | Yes | Yes | Yes | Include |
| Babiloni | 2021b | Yes | Yes | Yes | Yes | Yes | Yes | Yes | Include |
| Babiloni | 2022 | Yes | Yes | Yes | Yes | Yes | Yes | Yes | Include |
| Bender | 2014 | No | Unclear | Yes | Unclear | No | Yes | Yes | exclude |
| Birba | 2022 | Yes | Yes | Yes | Yes | Yes | Yes | Yes | Include |
| Bobes | 2010 | No | Yes | Yes | No | Yes | Yes | Yes | Include |
| Bonanni | 2008 | Yes | Yes | Yes | Yes | Yes | Yes | Yes | Include |
| Bonanni | 2010 | Yes | Yes | Yes | Yes | Yes | Yes | Yes | Include |
| Bonanni | 2021 | Yes | Yes | Yes | Yes | Yes | Yes | Yes | Include |
| Briels | 2020 | Yes | Yes | Yes | No | Yes | Yes | Yes | Include |
| Cao | 2022 | Yes | Yes | Yes | Yes | Yes | Yes | Yes | Include |
| Cecchetti | 2021 | Yes | Yes | Yes | Yes | Yes | Yes | Yes | Include |
| Dauwan | 2016a | DATASET VAN DELLEN 2015 | Yes | Yes | Yes | Yes | Yes | Yes | Include |
| Dauwan | 2016b | No | Yes | Yes | Yes | Yes | Yes | Yes | Include |
| Diaz-Rivera | 2023 | No | Yes | Yes | Yes | Yes | Yes | Yes | Include |
| Durongbhan | 2019 | No | Yes | Yes | Unclear | No | Yes | Yes | Include |
| Franciotti | 2020 | No | Yes | Yes | Yes | Yes | Yes | Yes | Include |
| Franciotti | 2022 | No | Yes | Yes | Yes | Yes | Yes | Yes | Include |
| Gaubert | 2019 | No | Yes | Yes | Yes | Yes | Yes | Yes | Include |
| Golob | 2009 | No | Yes | Yes | Yes | Yes | Yes | Yes | Include |
| Herzog | 2022 | No | Yes | Yes | No | Yes | Yes | Yes | Include |
| Jennings | 2022 | dataset Peraza, 2018 | Yes | Yes | Yes | Yes | Yes | Yes | Include |
| Kai | 2005 | Yes | Yes | Yes | Unclear | Yes | Yes | Yes | Include |
| Kim | 2021 | Yes | Yes | Yes | Unclear | Yes | Yes | Yes | Include |
| Kramberger | 2013 | Yes | Unclear | No | No | Yes | Yes | Yes | exclude |
| Lamos | 2021 | Yes | Yes | Yes | Yes | No | Yes | Yes | Include |
| Leko | 2018 | No | Yes | Yes | No | Yes | Yes | Yes | Include |
| Lian | 2021 | No | Yes | Yes | Yes | Yes | Yes | Yes | Include |
| Lindau | 2003 | Yes | Yes | Yes | Yes | Yes | Yes | Yes | Include |
| Massa | 2020 | Yes | Yes | Yes | Yes | Yes | Yes | Yes | Include |
| Maturana-Candelas | 2019 | Yes | Yes | Yes | Unclear | Yes | Yes | Yes | Include |
| Maturana-Candelas | 2020 | dataset Maturana-Candelas, 2019 | Yes | Yes | Unclear | Yes | Yes | Yes | Include |
| Mehraram | 2021 | No | Yes | Yes | Yes | Yes | Yes | Yes | Include |
| Moguilner | 2022 | No | Yes | Yes | Yes | Yes | Yes | Yes | Include |
| Moral-Rubio | 2021 | No | Yes | No | Unclear | Yes | Yes | Yes | Include |
| Musaeus | 2021 | Yes | Yes | No | Yes | Yes | Yes | Yes | Include |
| Nishida | 2011 | Yes | Yes | Yes | Yes | Yes | Yes | Yes | Include |
| Nishida | 2013 | Nishida, 2011 | Yes | Yes | Yes | Yes | Yes | Yes | Include |
| Ochoa | 2017 | Yes | Yes | Yes | Yes | Yes | Yes | Yes | Include |
| Pascarelli | 2020 | Yes | Yes | Yes | Yes | Yes | Yes | Yes | Include |
| Peraza | 2018 | No | Yes | Yes | Yes | Yes | Yes | Yes | Include |
| Perez-Valero | 2022 | No | Yes | Yes | Unclear | Yes | Yes | Yes | Include |
| Perez-Valero | 2022b | No | Yes | Yes | Unclear | Yes | Yes | Yes | Include |
| Pietto | 2016 | Yes | Yes | Yes | Yes | Yes | Yes | Yes | Include |
| Polverino | 2022 | Yes | Yes | Yes | Yes | Yes | Yes | Yes | Include |
| Quiroz | 2011 | Yes | Yes | Yes | Yes | Yes | Yes | Yes | Include |
| Revilla-Vallejo | 2021 | No | Yes | Yes | No | Yes | Yes | Yes | Include |
| Rochart | 2020 | Yes | No | Yes | Unclear | No | Yes | Yes | Include |
| Rodriguez | 2014 | Yes | Yes | Yes | No | Yes | Yes | Yes | Include |
| Rosenblum | 2020 | Yes | Yes | No | Unclear | Yes | Yes | Yes | Include |
| Rosenblum | 2022 | Yes | Yes | Yes | Yes | Yes | Yes | Yes | Include |
| Schumacher | 2019 | Yes | Yes | Yes | Yes | Yes | Yes | Yes | Include |
| Schumacher | 2020 | No | Yes | Yes | No | Yes | Yes | Yes | Include |
| Schumacher | 2021 | No | Yes | Yes | No | Yes | Yes | Yes | Include |
| Shim | 2022 | Yes | Yes | Yes | Yes | Yes | Yes | Yes | Include |
| Smailovic | 2017 | Yes | Yes | Yes | Unclear | Yes | Yes | Yes | Include |
| Smailovic | 2019 | Yes | Yes | Yes | Unclear | Yes | Yes | Yes | Include |
| Stothart | 2021 | Yes | Yes | Yes | Yes | Yes | Yes | Yes | Include |
| Stylianou | 2018 | Yes | Yes | Yes | Yes | Yes | Yes | Yes | Include |
| Suarez-Revelo | 2016 | Yes | Yes | Yes | Yes | Yes | Yes | Yes | Include |
| Tautvydaitė | 2022 | Yes | Yes | Yes | No | Yes | Yes | Yes | Include |
| van Dellen | 2015 | Yes | Yes | Yes | Yes | Yes | Yes | Yes | Include |
| Yu | 2016 | Yes | Yes | Yes | Yes | Yes | Yes | Yes | Include |
| Zhang | 2021 | No | Yes | Yes | Yes | Yes | Yes | Yes | include |
| Zhao | 2019 | Yes | Yes | Yes | Yes | Yes | Yes | Yes | include |

**S3**

|  | **First author** | **Year** | **Study population** | **EEG acquisition** | **Analysis** | **Results** |
| --- | --- | --- | --- | --- | --- | --- |
| 1 | Babiloni | 2017 | • 75 HC • 75 prodAD | • 19; 10-20; 128 Hz • rsEEG; 5min; EC; EOG | • FFT  • TF; IAF • eLORETA: ROI | • mean TF: PRODAD<HC • Posterior α2 and α3: PRODAD<HC • Parietal δ sources: PRODAD>HC |
| 2 | Babiloni | 2021b | • 60 HC • 63 prodAD | • 19; 10-20; 128Hz • rsEEG; EC; 3-5 min | • FFT  • α TF, IAF  • eLORETA | • age reduced rsEEG α in HC, effect reversed in PRODAD  • mean TF and IAF: PRODAD<HC |
| 3 | Babiloni | 2021a | • 70 prodAD • 60 HC | • 19; 10-20; 128-512Hz • rsEEG; 3-5min | • FFT  • TF (θ ; α), IAF  • eLORETA | • mean TF and IAF: PRODAD<HC  • HC EDU+: greater and widespread α activations • widespread α activations: PRODAD EDU+ < PRODAD Edu– |
| 4 | Briels | 2020 | • 197 SCD (A-/T-); 214 demAD (A/T) • 202 SCD (A-/T-/N-); 196 demAD | • 21; 10-20; 500Hz • 20min; rsEEG; EC | • Coh, iCoh, PLV, AEC, AEC-c, PLI, wPLI • FFT: mean global relative power | • demAD: AEC-c, AEC, coherence decrease in α and β; PLV decrease in α; iCoh, PLI and wPLI increase in θ |
| 5 | Cao | 2022 | • 19 demAD • 20 HC | • 128; 10-10; 2kHz • 30 min rsEEG EO+EC | • CWT, revisedHHT  • SVM; 5-fold cv | • RHHT: 86% CA in EO, 85% in EC • CWT: 78% CA |
| 6 | Cecchetti | 2021 | • 33HC • 39 demAD • 51 prodAD; 35 MCI | • 19 • rsEEG | • current density  • graph analysis | • demAD widespread slowing  • θ density: prodAD> MCI, HC  • β2 density(parietal+occipital): prodAD<HC |
| 7 | Durongbhan | 2019 | • 10 yHC (<70); 10 oHC (>70) • 20 AD | • 128; 10-10; 2000 Hz • rsEEG; 30 min (5min EO; 5 min EC) | • FFT; CWT • KNN; n-1 CV | • <70: sensitivity (>89% for FFT; >90% for CWT) specificity (>81% for FFT and >83% for CWT) • > 70: sensitivity (>64% for FFT; >67% for CWT) specificity (>84% for FFT and >85% for CWT) |
| 8 | Leko | 2018 | • 49 demAD • 28 prodAD • 4 HC | • 32; 10-20 • auditory oddball | • P300 • N200 | • P300 and N200 latency correlate to p-tau181, p-tau199 and ptau231  • N200 latency negatively correlated to Aβ1-42 |
| 9 | Lian | 2021 | • 43HC • 43 AD (CSF) | • 64; 10-20; 1000Hz • 5 min rsEEG EC | • microstate analysis • GFP • GEV | • no sign differences in average GEV and number of GP peaks • microstate duration, occurrence and coverage altered • Duration B: demAD>HC |
| 10 | Maturana-Candelas | 2019 | • 51 HC • 51 prodAD • 51 mild demAD, 50modeate demAD, 50 severe AD | • 19; 10-20; 500Hz • 5min; rsEEG; EC | • MSE; rMSSE • SMR; LOOCv; QDA; LDA |  |
| 11 | Maturana-Candelas | 2020 | • 51 HC • 51 prodAD • 51 mild demAD, 50moderate demAD, 50 severe AD | • 19; 10-20; 500Hz • 5min; rsEEG; EC | • BispRP, BispEn, BispMF | • BispRP: increases in δ and θ with disease-severity; decreases in α, β1, and β2 • BispEn: decrease δ and θ with severity, overall decrease • BispMF: overall decrease with AD severity |
| 12 | Perez-Valero | 2022 | • 8 demAD • 5non-AD MCI • 8HC | • 16; 10-20; 256Hz  • rsEEG; EO; 6min | • Relative power, SE, HC • selfdriven feature extraction and classification | • demAD vs HC: .91 precision, .82 recall, 0.88 F1 |
| 13 | Perez-Valero | 2022b | • 8 HC • 5 non AD-MCI • 8 demAD | • 16; 10-20; 256 Hz  • rsEEG; EO; 3min | • Relative power; HC; SE • SVM; LR; LOSO-CV | • demAD vs HC: .83 precision, .88 recall, .86 F1 |
| 14 | Pietto | 2016 | • 10 HC • 10 pFAD (E280A) | • 64; 500 Hz • ERP visual short term memory | • Monte Carlo; non-parametric bootstrapping • ROI • N1 (170ms), P2 (150-300ms), P3 (300-600ms), LPP (400-1000ms) | • N1, P2, P3: pFAD<HC |
| 15 | Polverino | 2022 | • 15 prodAD • 10 HC | • 256; 256Hz • rsEEG; EC; 20 min | • PSD | • α2, α2/ α1 ratio: prodAD<HC |
| 16 | Revilla-Vallejo | 2021 | •45 HC • 69 prodAD • 81 AD | • 19; 10-20; 200Hz • rsEEG; EC; 5min | • eLORETA • AEC • SE | • SE: δ band: AD> HC, MCI> HC; α: AD<MCI, AD<HC, β1: AD<HC; MCI<HC |
| 17 | Smailovic | 2018 | • 197 demAD • 230 prodAD • 210 SCD (A/T) | • 20s EC | • FFT • GFP, GFS | • ProdAD: low Aβ42 associated with high GFP δ and θ; high p-tau and t-tau associated with low GFP α and β • AD: high p-tau and t-tau associated with low GFP β and δ • AD, MCI: low Aβ42 low α GFS  • MCI: high p-tau and t-tau associated with low α GFS |
| 18 | Smailovic | 2019 | • 308 HC • 210 SCD (A/T) • 230 prodAD  • 197AD | • 19; 10-20; 256Hz • rsEEG; EC | • microstate analysis: k-means cluster analysis | • HC vs AD, SCD, MCI: topographic differences A, D  • prodAD, SCD vs AD: topographical differences A  • SCD vs AD, MCI, HC: topographic differences C  • Aβ42 linked to C; p-tau linked to B |
| 19 | Stothart | 2021 | • 20 yHC • 20 oHC • 20 AD | • 65; 400; 1kHz  • ERP recognition memory | • FFT  • SNR | • recognition response: AD<HC |
| 20 | Tautvydaitė | 2022 | • 29 demAD  • 19 HC | • 128; 512Hz • ERP encoding+ recognition | • ROI  • LORETA | • Medial temporal lobe activation New and N-back stimuli: HC> AD  • P2 New+N-back: HC>AD  • P300: repetition: AD>HC  • AD: no activation differences brain regions between New, N-back and repetitions |
| 21 | Zhang | 2021 | • 30 HC • 30 AD (A) | • 8; 10-20; 1000Hz • 5min | • Relative Spectral power • SpecEn • PSI | • Spectral power, SpecEn α: AD<HC  • Spectral θ: AD>HC • Spec En β: AD>HC • PSI δ, β, θ: AD < HC |
| 22 | Zhao | 2019 | • 40 HC • 40 AD | • 128; 10-10; 2kHz • 30min rsEEG EO-EC | • Linear+nonlinear FC: mean+RMS  • KNN | • CA > 70 (linear+nonlinear): EO 86.5%, EC 90.5%  •CA < 70 (lin+nonlin): EO 80.3%, EC 74.5% |

|  | **First author** | **Year** | **Study population** | **EEG acquisition** | **Analysis** | **Results** |
| --- | --- | --- | --- | --- | --- | --- |
| 1 | Arakaki | 2019 | • 10 HC  • 14 HC-PAT (Aβ/ T ratio) | • 21; 10-20  • WM; rsEEG | • SpecEn  • Α ERD | • low load: frontal α SpecEn CH-PAT>CH-NAT, decreased α ERD (nakijken) |
| 2 | Arakaki | 2022 | • 20 HC  • 21 HC-PAT (Aβ/ T ratio) | • 21; 10-20; 300Hz  • Stroop  • rsEEG; EO-EC 5min | • FFT; MWC  • ERD α (8–15 Hz)  • SpecEn | • low load: occipital α ERD low load: CH-PAT<CH-NAT (AUC=0.69); α SpecEn: CH-PAT> CH-NAT: frontal (AUC=0.73), occipital (AUC=0.69)  • low to high load frontal α SpecEn: CH-PAT< CH-NAT (AUC=0.72)  • low load late δ: HC-PAT<HC-NAT  • high load late θ : CH-PAT < CH-NAT |
| 3 | Bobes | 2010 | • 25 FAD (E280A) • 24 Acr (E280A) • 27 noncarriers | • 19; 10-20; 200Hz • ERP; semantic judgement | • N400: (311–490 ms) • IP • Intracranial source analysis | • AD, Acr: N400 generator strength anatomical shift • N400 amplitude: AD < acr |
| 4 | Gaubert | 2019 | • 175 HC  • 51 A-N+  • 63 preAD (A+N-); 25 preAD (A+N+) | • 256; 250Hz  • 1min; 30s EO-EC | • PSD in δ (1–4 Hz), θ (4–8 Hz), α (8–12 Hz), β (12–30 Hz), γ (30–45 Hz), MSF, SpecEn, algorithmic complexity, wSMI  in θ and α | • A-N+: decrease δ (frontocentral + temporal); higher β, complexity, SpecEn and wSMI θ ( frontocentral), higher γ (frontocentral+ temporal), increase MSF  • A+N+: increase γ (frontotemporal)  • A+N-: increase wSMI α (parieto-occipital) |
| 5 | Golob | 2009 | • 15 FAD Acr (PSEN1, APP) • 9 FAD noncarriers, 2HC | • 10; 10-20; 500Hz • auditory oddball (target detection) • A1+A2 | • stepwise dicriminant function analysis | • FAD Acr: delays latencies N100, P200, P300 • FAD Acr larger P200 amplitudes • P200 nontarget latency: 87% FAD classification |
| 6 | Kim | 2021 | • 24 SCD+ (A+); 146 SCD- • 29 prodAD (A+), 34 MCI- | • 19; 10-20; 200-250 Hz  • rsEEG; 3min  • A1-A2 | • PSD: FFT  • SVM | • SCD+ vs SCD- |
|  |  |  |  |  |  | • prodAD vs MCI- |
| 7 | Rochart | 2020 | • 10 HC • 14 preAD (A+T+) | • 21; 10-20; 300Hz • WM task | • TF: FFT, MWC, iFFT | • 0-back: low γ frontal+central: CH-PAT> CH-NAT • 2-back: low γ left temporal: CH-PAT<CH-NAT; high γ parietal: CH-PAT<CH-NAT |
| 8 | Shim | 2022 | • 69 SCD (A-)  •26 SCD (A+) | • 19; 10-20; 200-250Hz  • rsEEG | • PSD: FFT  • TAR, DAR, TBR  • sLORETA  • DMN analysis | • RP δ frontal, parietal and occipital: A+>A-  • frontal DAR: A+>A- |
| 9 | Suarez-Revelo | 2016 | • 15 PSEN1 Acr • 12 pFAD • 27 noncarriers | • 64; 10-10; 1000Hz | dDTF | • Occipito-parietal region 500-600 ms: Acr vs HC 67 |
|  |  |  |  |  |  | • Occipito-parietal region 500-600 ms: pFAD vs HC |
| 10 | Ochoa | 2017 | • 15 PSEN1 Acr • 15 non carriers | • 64; 10-10; 1000Hz • 5min; rsEEG; EC; tb memory encoding | • Transfer Entropy | • average connectivity: Acr> noncarriers |
| 11 | Quiroz | 2011 | • 10 Acr (PSEN1)  • 11 noncarriers | • 128 Hz  • Visual ERP | • 200-300ms | • 200-300ms: frontal positivity: carriers<HC, occipital positivity: carriers>HC |
| 12 | Rodriguez | 2014 | • 18 FAD (PSEN1) • 21 Acr  • 18 HC | • 19; 10-20; 200 Hz • rsEEG; EC | • AP, RP, MF, IAF • D2 • δ (0.5–3.5Hz), θ (3.5–7.5Hz), α (7.5–12.5Hz), β (12.5–19.1Hz) • ROC | • Global D2: HC vs Acr (ROC 0.90), HC vs AD (ROC 0.98), ACr vs AD (ROC = 0.92) • Regional D2: best accuracy ACr vs AD groups at the temporal region (ROC = 0.94) • Frequency 𝐷2: best accuracy β band: HC and ACr (ROC = 0.89), probable AD and HC (ROC= 0.99)  • decrease order of magnitude: HC vs AD (ROC = 0.98), ACr vs AD (ROC = 0.91), HC vs ACr (ROC = 0.89) |

|  | **First author** | **Year** | **Study population** | **EEG acquisition** | **Analysis** | **Results** |
| --- | --- | --- | --- | --- | --- | --- |
| 1 | Birba | 2022 | • 15 bvFTD  •15 AD • 21 HC | • 128; 1024Hz  • 10 min; rsEEG | • wSMI • nonparametric cluster-based permutation | •bvFTD: right frontotemporal hypoconnectivity |
| 2 | Diaz-Rivera | 2023 | • 14 bvFTD  • 30 HC  • 18 AD | • 128; 1024Hz  • Go/No Go; 15min | • SP • Polarity index | • bvFTD vs HC: differences in θ modulation; differences No-Go polarity index (δ) |
| 3 | Herzog | 2022 | • 99HC • 25 bvFTD • 49 AD | • 128; 1024 Hz • rsEEG; EC; 5min | • sLORETA  • HOFC • GSA • RF classifier | • δ : 0.96 AUC bvFTD vs HC |
| 4 | Lindau | 2003 | • 19 HC • 16 AD  • 19 FTD | • 10-20; 256Hz  • rsEEG; EC | • FFT dipole approximation • GFP, spectral ratio | •  α , ß1, ß2 and ß3: FTD<HC  • no differences δ or θ  • β1: 79% CA FTD vs HC |
| 5 | Moguilner | 2022 | • 76AD  • 54 bvFTD • 152HC | • 128; 1024Hz • rsEEG; 10min | • PLV; wSMI • k-fold validation, RF classifier, XGBoost classification | • bvFTD vs HC: AUC 0.78 |
| 6 | Moral-Rubio | 2021 | • 18 nfvPPA, 10 svPPA, 12 lvPPA  • 20 HC | • 32, 10-20 • 20min; rsEEG; EC | • DFT; WT • AutoEncoders • GTA • Decision Tree, EN, SVM, RF, kNN, Gaussian NB, multinomial NB | • PPA vs HC: kNN  • discrimination between PPA variants: kNN |
| 7 | Musaeus | 2021 | • 5 symptomatic FTD (CHMP2B) • 5 Acr • 6 non-carriers | • 18; 200Hz • alternating 30 s EC-EO | • Relative Power • GFP; GEV • ANCOVA | • Increased microstate D activity in duration <1 year, decrease in duration >2 years |
| 8 | Nishida | 2011 | • 19 FTD • 19 AD  • 22 HC | • 19; 10-20; 128Hz • rsEEG; EC; 15-20min | • GFP  •sLORETA | • α1 (orbitofrontal+ temporal) : FTD < HC : AUC 0.69 |
| 9 | Nishida | 2013 | • 19 FTD  • 19 demAD  • 19OHC, 18YHC | • 19; 10-20, 128Hz • rsEEG; 20min | • microstate analysis • GBF: k-means clustering | • Duration microstate C: FTD < HC • reversed sequence of activation C and D in FTD vs HC |

**Note.** Population in red is not biomarkerproven and was not included in the results.

|  | **First author** | **Year** | **Study population** | **EEG acquisition** | **Analysis** | **Results** |
| --- | --- | --- | --- | --- | --- | --- |
| 1 | Babiloni | 2018b | •30 prodAD •23 prodDLB • 30HC | • 19; 10-20; 128Hz • rsEEG, >5min | • FFT • TF (3-8Hz), IAF (6-14Hz) • eLORETA: LLC | • mean TF and mean IAF: HC>PRODAD>PRODDLB • HC vs prodAD: occipital α2  • HC vs prodDLB: parietal δ  • prodAD vs prodDLB: occipital α2 |
| 2 | Babiloni | 2019 | • 30 prodAD • 23 prodDLB • 30 HC | •19; 10-20; 128Hz • rsEEG; 5min EC | • TF; IAF • eLORETA; LCC | • inter- and intrahemispheric LLC α: prodAD=prodDLB< HC • HC vs PRODDLB intrahemispheric LCC global α2 AUROC 0.75  • HC vs prodAD interhemispheric LCC global α2 AUROC 0.72  • prodAD vs prodDLB: AUROC< 0.7 |
| 3 | Bonanni | 2021 | • 5 PPA, 13bvFTD  • 18 prodAD  • 20 HC | •19; 10-20; 256 Hz • rsEEG EC; 10 min | • MI | • pFTD vs HC: MI local left anterior > 0.30 (AUC 0.94); MI local right anterior >0.44 (AUC 0.83)  • prodAD vs HC: MI left local posterior > 0.30 (AUC 0.94); MI right local posterior > 0.30 (AUC 0.96)  • pFTD vs prodAD: MI left local anterior > 0.31 (AUC 0.71) |
| 4 | Dauwan | 2016b | • 66 DLB  • 66 prodAD  • 66 SCD | • 10-20 • 20 min rsEEG; EC | • PTE • MST • RF | • DLB vs AD: qEEG (highest β most important) • DLB vs HC: qEEG (θ/α ratio most important) |
| 5 | Franciotti | 2022 | Dataset Bonanni (2021) | • 19  • 10 min rsEEG; EC | • MI • graph theory analysis | • main hub of HC lost in onset FTD, substituted by provincial frontal hubs  • local efficiency Fp2, F4: FTD > HC; Local efficiency Fp2, C3: prodromal FTD>HC  • No changes in global network organization in AD |
| 6 | Massa | 2020 | • 12 prodDLB • 11 prodAD • 24 HC | • 19; 10-20 • rsEEG EC–EO; 20min | • FFT • PSA | • α/θ ratio: prodDLB< prodAD=HC |
| 7 | Yu | 2016 | • 48 bvFTD  • 69 AD  • 64 HC | • 21; 10-20; 500Hz • 20 min rsEEG | • PLI • MST | • δ PLI: bvFTD>AD • parietal α PLI: AD<bvFTD<SCD  • θ PLI: AD>FTD • differences in MST topology between AD, bvFTD and SCD in α band |
| 8 | Schumacher | 2020 | • 39 prodDLB • 36 prodAD  • 31 HC | • 128; 10-5; 1024Hz • EC rsEEG; 300s | • PSD: mean power; DF (4-15Hz) |  |
| 9 | Schumacher | 2021 | •37 prodDLB • 34 prodAD • 31 HC | •300s rsEEG EC, 30s EO •128 | • PSD: mean power, DF(4-15Hz) • α reactivity | •slowing in prodDLB vs HC and prodAD • α reactivity: prodAD=prodDLB < HC |

|  | **First author** | **Year** | **Study population** | **EEG acquisition** | **Analysis** | **Results** |
| --- | --- | --- | --- | --- | --- | --- |
| 1 | Aoki | 2019 | • 41 DLB • 80 HC | • 19; 500Hz • rsEEG; EC; 20 min | • ELoreta; ICA • RSN | • decreased occipital visual and sensorimotor network activity  • decreased occipital α |
| 2 | Babiloni | 2022 | •42 DLB  • 48ADD (McKhan, 2011) • 28HC | • 30; 10-20; 500-1024Hz • rsEEG; 3-5min EC, 3-5min EO | • FFT • BGF • power density • eLORETA | • reactivity of background posterior α: HC vs DLB  •mean TF, mean BGF: α-reactive HC > α-reactive DLB |
| 3 | Babiloni | 2018a | • 42ADD • 40HC •34DLB | • 19; 128Hz • rsEEG; 5min | • FFT • TF (3-8Hz), IAF (6-14Hz) • eLORETA: LLC | • mean TF, mean IAF: HC> DLB • Intra- and interhemispheric LCC in α: HC>DLB  • Interhemispheric LLC α2: 0.78 AUROC |
| 4 | Bonanni | 2008 | • AD  • 36 DLB  • 50 HC | • 19; 10-20, 1024Hz  • rsEEG, EC, 30min | • spectral power  • CSA; DF; DFV | • DLB: higher pre-α  • DLB vs HC: differences α-θ and α-δ ratios |
| 5 | Bonanni | 2010 | • 33AD • 26DLB  • 46HC | • 19; 10-20 • rsEEG EC, 30min; auditory oddball | • P300 | • P300 amplitude: delayed and lower amplitude in DLB  • P300 latency gradient inversion in DLB |
| 6 | Dauwan | 2016 | • 66AD  • 66 probable DLB  • 66 HC | • 21; 10-20; 500Hz  • 20 min rsEEG | • Spectral power  • dPTE | • DLB: dPTE gradient shift from posterior towards anterior in α band, loss of dPTE gradient |
| 7 | Franciotti | 2020 | • 111 AD • 63 HC • 137 DLB | • 12; 10-20;  • EC, EO every 2 min; 30 min | • FFT  • DF, DFV | • DLB mean DF decrease in anterior derivations: pre-α |
| 8 | Jennings | 2022 | • 12 AD  • 21DLB  • 15HC | • 128; 10-5; 1024hHz • EC-EO; 150s | • DF, DFV • NCA • loocv, 10-fold CV • k-nn; SVM; LR | • DLB: decrease DF, decrease θ-α relative power |
| 9 | Kai | 2005 | • 15 AD • 15DLB  • 12 HC | • 14; 10-20; 1000Hz | • spectral power, Icoh | • DLB: increase δ, θ power • DLB: lower Icoh and Hcoh δ, θ, α, and β |
| 10 | Lamos | 2021 | • 21 prodDLB  • 21 HC | • 256 • 5min; rsEEG; EC | • microstate analysis: individual local maxima of GFP • group-level clustering, k-means • IAF | • B significant shorter mean duration and increased occurrence in prodDLB • A-D higher occurrence in prodDLB •prodDLB lower IAF |
| 11 | Mehraram | 2021 | • 18HC • 32AD  • 25DLB | • 128; 10-5; 1024Hz • rsEEG, 2,5min | • WPLI  • random forest; 6-fold CV | • α WPLI: DLB<HC • DLB vs HC: WPLI β band, modularity, characteristic path length and clustering coefficient α |
| 12 | Pascarelli | 2020 | • 60DLB  • 60 AD | • 256-1024Hz; 19; 10-20 • rsEEG > 5min | • IAF, TF • eLORETA | • mean TF and mean IAF: HC, AD > DLB • DLB decrease of eLORETA solutions in posterior α2 and α3 sources • DLB VH+ vs HC: parietal δ source activity |
| 13 | Peraza | 2018 | • 17 HC • 26AD •25DLB | •128; 10-5; 1024Hz  •150srsEEG EC | • DF • PLI • MST | • DF (occipital): DLB < HC  • PLI α: DLB<HC  • MST: more randomization |
| 14 | Rosenblum | 2020 | •23 DLB • 22 HC | • 19; 10-20; 500 Hz  • auditory+visual oddball | • ERO: P3  • ITPC •MWC | • θ ERO (visual): DLB<HC  • α, β: DLB>HC  • δ ERO: DLB<HC  • P300 latency: DLB>HC |
| 15 | Rosenblum | 2022 | • 28 DLB  • 32 HC | • 19; 10-20; 500Hz • Visual oddball | •TF: Morlet Wavelet Convolution • ROC/ AUC | • Δ target ERS: DLB < HC • Β suppression: DLB<HC  • α suppression: DLB-VH+ < DLB-VH− • DLB vs HC: δ (75% sens, 72% spec), θ(71% sens, 69% spec), α (57% sens,56% spec), β (71% sens and 63% spec) |
| 16 | Schumacher | 2019 | • 25 DLB  • 27 AD  • 18HC | • rsEEG (EC) •128; 10-5; 1024 Hz | • microstate analysis | • Microstate duration: DLB > HC • Distinct microstates per second: DLB < HC |
| 17 | Stylianou | 2018 | • 18 AD  • 17 DLB  • 21HC | • 128; 10-5; 1024Hz • rsEEG, EC | • DF, FP, DFV | • θ: DLB>HC; β: DLB<HC; α: DLB<HC • θ-α DF and α DF: DLB<HC  • HC: lower FP slow-θ range than DLB in all regions |
| 18 | Van Dellen | 2015 | • 66 DLB  • 66 AD  • 66HC | • 21; 10-20; 500Hz • rsEEG EC | • FFT • PLI, MST | • MST: DLB < HC  • PLI α: DLB < HC |
